# The Role of Community Beliefs and Practices on the Spread of Ebola in Uganda, September 2022

**DOI:** 10.1101/2023.12.05.23299506

**Authors:** Helen Nelly Naiga, Jane Frances Zalwango, Saudah N. Kizito, Brian Agaba, Brenda N. Simbwa, Maria Goretti Zalwango, Rebecca Akunzirwe, Zainah Kabami, Peter Chris Kawugenzi, Robert Zavuga, Mackline Ninsiima, Patrick King, Mercy Wendy Wanyana, Thomas Kiggundu, Richard Migisha, Doreen Gonahasa, Irene Kyamwine, Benon Kwesiga, Daniel Kadobera, Lilian Bulage, Alex Ario Riolexus, Sarah B. Paige, Julie R. Harris

## Abstract

**Background:** Traditional community beliefs and cultural practices can facilitate the spread of ebolaviruses during outbreaks. On September 20, 2022, Uganda declared a Sudan Virus Disease (SVD) outbreak after a case was confirmed in Mubende District. During September–November 2022, the outbreak spread to eight additional districts. We investigated the role of community beliefs and practices in the spread of SUDV in Uganda in 2022.

**Methods:** A qualitative study was conducted in Mubende, Kassanda, and Kyegegwa districts in February 2023. We conducted nine focus group discussions (FGDs) and six key informant interviews (KIIs). FGDs included SVD survivors, household members of SVD patients, traditional healers, religious leaders, and community leaders. Key informants included community, political, and religious leaders, traditional healers, and health workers. We asked about community beliefs and practices to understand if and how they contributed to the spread of SUDV. Interviews were recorded, translated, transcribed, and analyzed thematically.

**Results:** Frequently-reported themes included beliefs that the community deaths, later found to be due to SVD, were the result of witchcraft or poisoning. Key informants reported that SVD patients frequently consulted traditional healers or spiritual leaders before seeking formal healthcare or visited them after formal healthcare failed to improve their health conditions. They also noted that traditional healers treated patients with signs and symptoms of SVD without protective measures. Additional themes included religious leaders conducting laying-on-of-hands prayers for SVD patients and symptomatic contacts, SVD patients and their symptomatic contacts hiding in friends’ homes, and exhumation of SVD patients originally buried in safe and dignified burials, to enable traditional burials.

**Conclusion:** Varied community beliefs and cultural practices likely promoted SVD outbreak spread during the 2022 outbreak in Uganda. Controlling ebolavirus outbreaks in Uganda could be aided by the involvement of formal public health systems, traditional healers, and religious leaders. Community engagement during inter-epidemic periods could aid in the effective management of future outbreaks in Uganda by identifying socially acceptable and scientifically supported alternatives for infection control.

## Introduction

Ebola disease (EBOD) is a viral hemorrhagic fever suspected to be transmitted from infected primates, bats, or other wildlife to humans, generally through human contact with blood or body fluids [1, 2]. Infection in humans usually causes acute fever and, in many people, a hemorrhagic syndrome later in the course of disease that is fatal in 50-90% of cases, even with treatment [3, 4]. Contact with the body fluids of infected patients can lead to infections in their household members, close contacts, and facility-based caregivers [5-8]. Even after the death of a patient, the body remains highly infectious, and contact with the bodies of persons who have died of Ebola without appropriate personal protective equipment is considered extremely high-risk [9].

In African settings, community members often seek advice from traditional healers or religious leaders before, or in place of, accessing formal medical care [10]. Practices among these persons can involve very close contact without appropriate protective equipment. An example is the treatment of “false teeth” (locally called *ebinyo*), which involves the gouging-out of unerupted teeth, believed by many communities to be a source of illness in children [10, 11]. Other practices include crude tonsillectomies to treat *gapfura*, a childhood illness usually characterized by respiratory disease, in which a child’s tonsils and throat are manually scraped by a traditional healer until blood and pus drain from the mouth [12]. Beyond traditional treatments, some cultural practices, such as washing and touching of the body of dead persons by mourners at a funeral, can also spread infection [13].

On September 20, 2022, for the first time in over a decade, Uganda declared an outbreak of Sudan Virus Disease (SVD) caused by the Sudan virus (formerly called *Sudan ebolavirus*). By the end of the outbreak, there were 142 confirmed cases and 22 probable cases from nine districts [14, 15]. Anecdotal reports from the epidemiologic investigations revealed that many patients had undergone treatment by traditional healers or religious leaders for their illnesses before they were diagnosed with SVD.

To effectively provide interventions to stop an outbreak, it is necessary to understand specific cultural practices and beliefs that may facilitate disease spread, and to work with the community to design interventions that can maintain safety while respecting important cultural aspects [1, 16]. However, the role of such beliefs and practices and how they may have contributed to the outbreak were poorly understood. We evaluated how community beliefs and cultural practices may have contributed to the spread of Ebola across Uganda, in 2022.

## Methods

### Study design and site

We conducted a qualitative study in Mubende, Kassanda, and Kyegegwa Districts. We conducted key informant interviews (KII) and focus group discussions (FGDs) with healthcare workers (HCW), village health team members (community health workers, also called VHTs), traditional healers, religious leaders, and district surveillance staff who participated in the response of the outbreak.

We also reviewed the case investigation reports collected by the field teams during the outbreak to summarize self-reported visits to traditional healers and religious leaders occurring after the onset of illness for patients.

#### Data collection

The study participants were identified, consented, recruited, and organized with the assistance of VHTs in their communities. All interviews were conducted in the local language (Luganda) and used interview guides.

We conducted nine FGDs, each with a maximum of 10 participants. Four FGDs in Mubende District, four FGDs in Kassanda District, and one FGD in Kyegegwa District. Only one FGD was conducted in Kyegegwa because, even though Kyegegewa borders Mubende and Kassanda, there were very few cases compared to Mubende and Kassanda districts. In the FGDs, we asked about the different community practices and beliefs that could have contributed to the spread of SVD in these districts. Each FGD included five males and five females. The participants were selected by convenience sampling by the village health team leaders (VHTs) from the affected villages.

Six KIIs were conducted. These included two health workers from the clinics where most early cases were treated, two traditional healers, and two religious leaders. Three KIIs were conducted in Mubende District and three KIIs in Kassanda District. We purposively selected the key informant participants from the most affected subcounties in the two districts.

### Data analysis

FGDs and KIIs were audio recorded, translated into English, and transcribed verbatim for analysis at the end of each data collection day. Data were stored by the principal investigator on a password-protected computer. Participants were identified based on the type of interview and district of residence (e.g., FGDM1 represents FGD Mubende 1; FGDK1 represents FGD Kassanda 1). This code was applied to transcripts. Transcripts were reviewed by the study investigators, coded, and analyzed thematically using the CDC Excel thematic analysis tool, ver. 10.18.22 [1]

### Ethics approval and consent to participate

The Ministry of Health of Uganda gave the directive and approval to carry out this investigation. Further approval to conduct this study was obtained from Mubende and Kassanda District offices, Mubende Regional Referral Hospital case management team, the National Institute of Public Health and Uganda Ministry of Health. This activity was reviewed by CDC and was conducted consistent with applicable federal law and CDC policy.§See e.g., 45 C.F.R. part 46, 21 C.F.R. part 56; 42 U.S.C. §241(d); 5 U.S.C. §552a; 44 U.S.C. §3501 et seq. Participants provided informed written consent before participating in the study.

## Results

Five major themes emerged potentially contributing to the spread of SVD among the different districts: 1) engaging traditional healers; (2) communalism; 3), touching ill persons during prayers; (4) attribution of illness to supernatural forces, and 5) cultural practices around burials. Reviews of case investigation reports identified at least 11 patients who went to one or more traditional healers during their illnesses and received unspecified treatment, and four who went to a traditional healer and had *ebinyo, gapfura*, or removal of the epiglottis performed. At least seven patients underwent traditional prayers for healing during which religious leaders laid hands on them to heal them. Two SVD patients were traditional healers and two were religious leaders.

### 1. Engaging traditional healers

FGD and KII participants reported that when someone falls sick suddenly, it is usually assumed to be due to witchcraft. At the beginning of the outbreak, many FGD participants in Mubende District felt that Ebola was a curse upon their community and that traditional medicine was a necessary intervention to counteract the curse. They believed that traditional medicine would reverse the witchcraft and thus reduce the community deaths. However, participants reported that “the traditional practices only spread Ebola to other areas”.

*‘‘…When the child started passing out blood from the mouth suddenly, we thought we needed to consult with the traditional healers because we all thought it was a traditional illness. We needed to consult our ancestors because her illness was strange. She was vomiting blood so we needed to know where the blood was coming from. We took her back to our home district. The traditional healer, her grandmother, and others got infected with Ebola.”*

*-FGD participant, female, Mubende*

*‘‘…In the beginning, many community members died, but we thought it was witchcraft and because many were passing out blood, rumors had it that it was poisoning. So the community members went to traditional healers to help them suck that poison out of their bodies using traditional herbs instead of seeking care in the health facilities. This contributed to the spread of Ebola in our community and the neighboring districts.”*

*-KII participant, female, Mubende*

*‘‘…when our brother fell sick, we thought our enemies had cast a spell on him, so we took him to a traditional healer in Luweero and another one in Kampala. After his death, we lost six other family members to Ebola and two survivors.”*

*-FGD participant, male, Mubende*

*‘‘… when my brother started coughing and vomiting blood, my mother suggested that we take him to Kampala to a renowned traditional healer. Taking my brother to (a traditional healer in) Kampala led to the first cluster of Ebola cases in Kampala.”*

*-FGD participant, male, Kassanda*

Participants reported that some traditional healers ended up contracting the disease themselves and passing it to their other patients or family members.

*‘‘*… *My mother was a traditional healer, (and) she treated a young girl who was vomiting and coughing blood from Mubende District. The girl’s mother thought she had false teeth but my mother had to check if her tonsils had burst. So, mother used her bare hands to check where the blood from this girl’s mouth was coming from. Days after she treated this girl, Mother fell sick. She was infected with Ebola and died a week after she was taken to the Ebola treatment unit in Mubende. My young brother and father also got Ebola but survived.”*

*FGD participant, female, Kyegegwa*

*‘‘*…*My wife treated one of the first patients in Kassanda with different herbs as the entire community initially thought he was passing out blood because of witchcraft. This young man was a thief of goats…so the community members thought he had been bewitched this time. My wife got infected with Ebola and died. My maid (who took care of my wife while she was sick) and I also got infected with Ebola but luckily survived.”*

*FGD participant, male, Kassanda*

### 2. Communalism

The FGD participants reported that if a community member is sick or has a patient, the entire community is obliged to support that member. The participants noted that the practice of shared responsibility also contributed to the spread of Ebola in this community and other areas.

*‘‘…In this community, people are always willing to help. I invited my neighbors to help me carry the child when she was vomiting blood. I didn’t know this was Ebola. This is how my neighbors got infected with Ebola”*.

*FGD participant, female, Mubende*

*‘‘… one of the first Ebola cases in Kassanda was a young man who had a butchery and roasted goat’s meat in the evenings. When he fell sick…his many friends came to visit him in the clinic. At the time of his evacuation, he was vomiting blood but many of his friends were there to support him. This communalism led to so many other Ebola cases and deaths in our community”*.

*FGD participant, male, Kassanda*

*‘‘…when our elder sister fell sick, we all participated in taking care of her until she passed on. After her death, we lost four other members of our family to Ebola and two Ebola survivors”*.

*FGD participant, female, Mubende*

*‘‘….[when] my brother started vomiting blood, we moved to Kampala, to live with my mother’s sister for better management. This led to all these family members, the neighbors and I catching Ebola in Kampala. We lost so many people to Ebola”*.

*FGD participant, male, Kassanda*

*‘‘…In our culture, it is an abomination for someone to fall sick, and fail or refuse to visit them while in the health facility. Many of these young men got infected during those moments because (an Ebola case) was heavily bleeding. Many took it upon themselves to participate in bathing and washing clothes for the patient. This exposed many of them to Ebola. It is very unfortunate that we lost most of these youth who would make our community a better place in the future”*.

*KII participant, female, Kassanda*

### 3. Touching ill persons during prayers

FGD participants reported that religious healers who prayed for Ebola victims by laying hands on the sick not only ended up contracting the disease themselves but passing the disease to other people. It was reported that different church members contracted Ebola virus after they participated in a healing prayer session in which they laid hands on the body of a sick person.

*‘‘…As a pastor, when praying for someone seeking spiritual healing, you can’t avoid getting in contact with them. We lay hands on them for a special healing and anointing. This exposed us as spiritual healers to Ebola but also our family and church members. We lost some church elders in the church (prayer warriors and intercessors) to Ebola. My family and I are also survivors of Ebola.”*

*FGD participant, male, Mubende*

*‘‘…during that period, many people were seeking for spiritual healing as (they) believed their community had been put under a spell. So my son, being a pastor in this community, prayed for so many sick people. This exposed him, his wife, and son to Ebola. My grandson died but his parents are Ebola survivors.”*

*FGD participant, female, Mubende*

*‘‘…Many sought spiritual healing in our church. My wife and I prayed for some neighbors who had malaria-like symptoms but later started having bleeding symptoms. My wife shortly fell ill, she was six months pregnant when she was taken at the Ebola treatment unit. I am glad she survived Ebola but sadly a few days after her discharge, we lost our baby.”*

*FGD participant, male, Mubende*

### 4. Attribution of illness to supernatural forces

FGD and KII participants reported that there was general lack of understanding of the Ebola disease, and this had a negative influence on adherence to preventive measures. Many community members initially believed that the illnesses were caused by witchcraft or were punishment from God.

*‘‘…We didn’t know what this disease was and how it came in our community. I thought it was a disease found on the borders of Congo. I had heard of that disease but in DRC and at its borders. That is why many people fell sick and others died when Ebola was in our district. Many of us thought this was witchcraft or a punishment from God.”*

*FGD participant, male, Kassanda*.

*‘‘*… *During the Ebola lockdown in our district, we didn’t practice the control measures because most of us as young men, we thought it was witchcraft (and) not Ebola. When our friends fell sick, we would visit them. This led to more Ebola cases in (the village). We only believed when our friends started dying. This scared us and we eventually started practicing the preventive measures.”*

*FGD participant, male, Kassanda*.

*‘‘*…*The community didn’t believe the cause of death among the many community deaths in early August was a medical disease. They believed it was a traditional spell, so many more people died as all the burials were never supervised”*.

*KI, male, Mubende*

*‘‘…Our communities also believed that the community members who died in the earlier days…were poisoned by some new community members to steal their land as rumor had it that our land had gold.”*

*FGD participant, male, Mubende*

*‘‘*…*A very old man (over 100 years) died, and his people started dying, the family lost six people in a duration of one month. The rumors had it that the old man wouldn’t die alone, he had to take some relatives with him as it was believed that he was a witch doctor.”*

*FGD participant, male, Mubende*

### 5. Religious and cultural practices around burials

In Uganda, funeral and burial practices hold extreme significance, as they are perceived as crucial steps in transitioning from the world of the living to the spiritual world. Common funeral rituals include washing and cleaning of the deceased person’s body before burial. If a person being buried fails to obtain the appropriate ancestral spiritual rituals - which should be given by the surviving close relatives – it is believed that the deceased’s spirit may return and punish the living relatives. As a result, some community members exhumed bodies of persons who had been buried in safe and dignified burials to achieve the appropriate rituals before re-burial.

*‘‘…one of the Ebola cases in Kassanda was from a very strong Moslem background. When he passed on, the health burial team buried him at night. However, his relatives decided to exhume his body to bury him in the right Islam (way) (wash his body), perform all the Islam funeral practices and confirm that all his body parts were intact”*.

*KI, male, Kassanda*

*‘‘*…*the two men who exhumed [an Ebola patient]’s body, bathed and wrapped this body, all got infected with Ebola and died. These two men were known for exhuming bodies and transferring the graves in case someone has sold their burial grounds. These two men were in their late 50s so they were not friends with [the patient]”*.

*FGD participant, female, Mubende*

*‘‘…In our culture, when someone dies, we have so many rituals that we practice. Some of which include bathing, shaving and rolling the dead body in different clothes. In the earlier days of August, as a community, we didn’t know that this disease was so contagious. That is how Ebola spread to other areas and killed many of our people”*.

*FGD participant, male, Mubende*

*‘‘*…*During burials of some community members who had been bleeding in their late stages of their illnesses, some people would come from other districts like Kampala. These people would be close relatives of the deceased. They actively participated in the burial rituals, this led to some Ebola cases in Kampala”*.

*KI, male, Mubende*

## Discussion

We identified multiple beliefs and practices that contributed to the spread of Sudan virus in Uganda during 2022. These included engaging traditional healers for treatment, communalism, touching ill persons during prayers, attribution of illness to supernatural forces, and conduct during traditional burials. Such practices should be recognized for their potential contribution to spread of infection during ebolavirus outbreaks in Uganda and similar settings. A follow up study to identify the acceptable alternatives to these approaches together with the community could help reduce spread of infection in future outbreaks.

Traditional healers are regarded as custodians of daily traditional and cultural values in many communities in Africa, and using them as the first line of care during an illness is common [18, 19]. Media reports from 2021 suggested that traditional healers outnumbered medical doctors by a ratio of 35:1 in Uganda [20]. Traditional healers are often more highly regarded than persons who promote conventional medicine or other forms of health care [18]. Many practices of traditional healers in Uganda put them – and subsequently their future patients – potentially at risk of ebolavirus infections during an outbreak. A 2003 study from Northern Uganda reported traditional healers making incisions into people’s bodies and rubbing in herbal medicine, which led to the spread of ebolavirus [21]. Reports for at least 15 patients in our study documented their visits to traditional healers while they were ill; participants in our qualitative study suggested that such visits led to at least some subsequent SUDV infections.

Indeed, at least two patients in the outbreak were themselves traditional healers. Because of the preference in some communities for traditional healers as the first line of treatment, some have advocated for the engagement of traditional healers in controlling the spread of ebolaviruses [4, 16, 18]. In a study in West Africa, traditional healers were trained about ebolaviruses, how to recognize symptoms early, where suspected patients should be referred, and how to protect themselves; they were subsequently involved in risk communication for their communities. The communities trusted their communication, ultimately supporting disease control in the outbreak [18]. Another study in northwestern Uganda that described the training of traditional healers to identify plague symptoms and refer patients noted the importance of not stigmatizing or treating traditional healers as ‘less than’ conventional health providers. The study emphasized the importance of respecting differences in belief and practice to facilitate collaborative relationships for epidemic control [2].

Some people believed that the illness caused by SDV infection was caused by supernatural beings, witchcraft from neighbors or vengeance from an offended god as a result of transgressions committed in the past by an individual or parents. This is commonly observed during ebolavirus outbreaks, which frequently occur in communities with a strong belief in witchcraft as a cause of illness [1, 2,9]. A study in Nigeria also found that community members, especially those living in rural areas, believed that most health problems were spiritually related and could be resolved without allopathic medicine, resulting in their seeking treatment from a traditional healer to remove the witchcraft rather than a hospital [22]. In Sierra Leone, both traditional healers and community members initially attributed outbreak-related deaths to witchcraft or poisoning [23].

Religious and spiritual healing practices (touching ill persons during prayers) may also have contributed to the spread of the 2022 SDV outbreak in Uganda. Multiple patients had documented experiences during their illness of visiting religious leaders and those healers themselves also represented some of the outbreak patients. Community members reported that they sought spiritual healing to dispel the ‘witchcraft’. Similar findings were reported in a study in West Africa, in which religious leaders both contracted the illness from sick persons and passed it to others in prayer sessions [18, 24]. A subsequent intervention engaging religious leaders enabled them to educate their institutional members about the importance of seeking healthcare first in health facilities, isolating if ill, and using protective equipment in case of unexplained bleeding [25].

The normal cultural practices around funerals in many African settings, which involve washing and touching the body by family and mourners, can contribute to Ebola viruse(or SDV) spread during an outbreak [26]. Such practices, according to community members, are highly valued because they are regarded as critical steps in the transition from the world of the living to the spiritual world, which must be facilitated by surviving relatives through specific rituals [2, 4, 13, 18]. Similar to other studies, we heard from participants that funeral practices contributed to the spread of SUDV in Uganda [14, 21, 27]. Other practices reported by study participants, such as exhumation, are related to the lack of acceptance of safe and dignified burials, and contributed to the spread of SUDV. The rapid burial of the deceased without allowing relatives to view the dead bodies gave rise to the suspicion that medical professionals were keeping the corpses in order to sell their relatives’ body parts [14]. During a SUDV outbreak in Northern Uganda in 2003, such mistrust was exacerbated by rumors that some Westerners were purchasing human body parts [8, 28]. Without mutual trust between community members and health workers, adherence to preventive measures during an Ebola outbreak is likely to be compromised. In some communities, health officials attempting to conduct safe and dignified burials were attacked and prevented from executing their duties [28]. Initiating community engagement studies to identify appropriate and acceptable ways to allow them to participate in safe and dignified burials could reduce illness spread in a future SVD outbreak.

## Conclusion

The recent Sudan virus outbreak response in Uganda has demonstrated the importance of involving community, traditional, and religious leaders to facilitate disease control during outbreaks. Collaboration between health workers and these leaders at all stages of outbreak response campaigns, especially when the interventions include activities incompatible with affected communities’ cultural and religious practices or beliefs, may help effectively contain disease spread.

## Data Availability

The datasets upon which our findings are based belong to the Uganda Public Health Fellowship Program. For confidentiality reasons, the datasets are not publicly available. However, the datasets can be made available upon reasonable request from the corresponding author and with permission from the Uganda Public Health Fellowship Program.

## Recommendations

Engagement of formal public health systems with traditional healers and religious leaders in Uganda may help facilitate ebolavirus outbreak control. Community engagement activities during inter-epidemic periods – before an outbreak happens - to identify alternative acceptable and scientifically-backed actions to control infection is likely to support disease control in future outbreaks. For example, holding community educational sessions on ebolavirus-related risks to traditional healers and the community and dialogues on acceptable practices around safe and dignified burials during a non-outbreak period may help communities feel that they are stakeholders in the response and contribute to buy-in when an outbreak does occur. It may also be useful to engage formal healthcare professionals with traditional and religious leaders’ regional and national associations to discuss these approaches and how to implement them in the event of an outbreak. Such approaches could also help bring these leaders on board when an outbreak of any kind occurs.

## List of abbreviations

EBOD: Ebola disease
FGD: Focus group discussion
KII: Key informant interview
TH: Traditional Healer
DSFP: District Surveillance focal person
SUVD: Sudan Virus
SVD: Sudan Virus Disease
EBOV: is Ebola Zaire Virus

## Declarations

### Ethics approval and consent to participate

The Ministry of Health of Uganda gave the directive and approval to carry out this investigation. Further approval to conduct this study was obtained from Mubende and Kassanda District offices, Mubende Regional Referral Hospital case management team, the National Institute of Public Health and Uganda Ministry of Health.

This activity was reviewed by CDC and was conducted consistent with applicable federal law and CDC policy.§ §See e.g., 45 C.F.R. part 46, 21 C.F.R. part 56; 42 U.S.C. §241(d); 5 U.S.C. §552a; 44 U.S.C. §3501 et seq.

“*Sarah Paige was employed by USAID when the research for the current paper was conducted*.

*The views and opinions expressed in this paper are those of the authors and not necessarily the views and opinions of the United States Agency for International Development*.”

## Consent for publication

Not applicable.

## Acknowledgments

The authors thank the staff of the Public Health Fellowship Program for the technical support and guidance offered during this study. The authors also extend their appreciation to Mubende and Kassanda district local council and health authorities for the support offered during the investigation.

## Copyright and licensing

All materials in the Uganda National Institute of Public Health Quarterly Epidemiological Bulletin is in the public domain and may be used and reprinted without permission; citation as to source; however, is appreciated. Any article can be re-printed or published. It should be referenced in the original form if cited as a reprint.

## Funding source and disclaimer

This outbreak investigation was supported by the President’s Emergency Plan for AIDS Relief (PEPFAR) through US Centers for Disease Control and Prevention Cooperative Agreement number GH001353–01 through Makerere University School of Public Health to the Uganda Public Health Fellowship Program, Ministry of Health. The contents of this article are exclusively the responsibility of the authors and do not essentially represent the official views of the US Centers for Disease Control and Prevention, the United States Agency for International Development, Makerere University School of Public Health, or the Ministry of Health, Uganda.

## Authors’ contributions

HNN took the lead in conceptualizing the study idea, data collection, data analysis, writing, and editing of the manuscript. JFZ, SNK, BA, GMZ, BNS, RA, ZK, PCK, RZ, MN, PK, MWW, and KT were involved in data collection, data analysis, and writing of the manuscript. RM, DG, IK, BK DK, LB, ARA, SP, and JH were involved in conceptualizing the study idea and writing, editing, and reviewing the manuscript. All authors read and approved the final manuscript.

